# THE RELATIONSHIP OF THE FOOT AND ANKLE STRUCTURE WITH OVERUSE INJURIES IN LICENSED FOOTBALLERS, A PROSPECTIVE COHORT

**DOI:** 10.1101/2020.07.22.20159764

**Authors:** Ali Sahillioglu, Lale Cerrahoglu

**Affiliations:** Department of Physical Medicine and Rehabilitation, Sisli Hamidiye Etfal Training and Research Hospital, Istanbul, Turkey; Department of Physical Medicine and Rehabilitation, Celal Bayar University Medical Scholl, Manisa, Turkey

**Keywords:** Ankle, Foot, Football, Joint Hypermobility, Overuse Injury

## Abstract

The main purpose of this study was to compare the values determined in the clinical examination of the foot and ankle and the plantar pressure measurements of the foot in athletes who developed an overuse-type disability and in athletes who did not develop overuse-type disability, and was to find factors that might predispose to disability, during the one-year follow-up. 100 licensed football players were included in the study. Presence of joint hypermobility, foot posture assessment, ankle and first metatarsophalangeal (MTP) joint range of motion (ROM) measurements, pedobarographic plantar pressure assessment of foot was carried out. Then, the footballers were followed for 12 months for the development of new foot and ankle overuse injuries and the clinical and pedobarographic data of the footballers with at least one injury were compared with the group without injury. We found asymmetric pressure distribution between the preferred and non-preferred foot in the group who had an injury in the pedobarographic static foot plantar pressure measurements *(p = .040)*. When we compared the ROM values of footballers who had an injury and footballers who did not have an injury, we found a significant limitation in the group that suffered an injury, in the ankle eversion, first MTP joint dorsiflexion and ankle plantarflexion degrees *(p = .029, p = .023, p=.044, respectively)*. These findings suggest that impairments in foot plantar pressure distribution and limitations in ankle and foot joint ROM may be risk factors for the development of foot and ankle overuse injury.

## 1. INTRODUCTION

Football is one of the most popular sport branches in the world with the spectator potential and the economy it creates. Game rules, tactical developments, and the quality of the football players increase the speed, intensity, and quality of the game. This situation has brought a competitive environment to football and accordingly the increase in the speed of the game has increased the number of double struggles and the severity of the game. Therefore, football has a high incidence of injury compared to other sports.^1^

Foot and ankle are some of the most frequently injured places in football players’ bodies.^2^ Disturbances in the structure and function of the foot can lead to prominent restrictions on the activity and performance level of the athlete. The full functioning of the foot is not only important in terms of foot health, but it is also necessary for the health of our entire body, especially neighboring joints.^1,3^

According to consensus statements, for football, overuse injury is defined as “one caused by repeated micro-trauma without a single, identifiable event responsible for the injury”.^4^ Overuse injuries have insidious onset and can restrain athletes from sports temporary or even permanently.^5,6^ Lower leg and in particular ankle and foot are highly involved in many sports and are vulnerable to overuse injuries. The most common types of foot and ankle overuse injuries are Achilles tendinopathy, metatarsalgia, metatarsal stress fracture and plantar fasciitis.^7^

It is thought that the risk factors that may cause injury can be shown by revealing the soles of the foot and the corrections to be made can contribute to the improvement of sports performance. Foot loading patterns in football have been described in previous studies, and alterations in biomechanical loads have been suggested to be closely associated with foot and ankle overuse injuries. Various foot and ankle problems result from repetitive high loads on the foot.^8–10^

Our aim was to compare the values determined in the clinical examination of the foot and ankle and in the plantar pressure measurements of the foot in athletes who developed overuse disability and in athletes who did not develop overuse disability. Thereby we aimed to identify the factors that might predispose to overuse injury with the data we obtained during the one-year follow-up.

## 2. MATERIAL AND METHODS

### 2.1 Subjects

Participants were evaluated at the Physical Medicine and Rehabilitation Polyclinic of Celal Bayar University Medical Faculty Hospital. A total of 104 football players, who played football in the A2 league, which is the league of the second teams of the professional teams, who regularly practice football in Izmir and Manisa provinces, were planned to be included in the study. 4 of the players did not fit the inclusion criteria. For this reason, a total of 100 licensed football players were included in the study. Age, height, weight, and body mass index (BMI) values of all the subjects were measured.

### 2.2 Selection Criteria of Study

Verbal and written information was given about the purpose of the work to be done and the nature of the practice to the participants. Participants who wanted to participate in the study were informed about the subject and individual approvals were obtained. It was determined that the weekly training program was on average for 3 days a week and 2 hours a day in each team. Weekly training programs were asked to the participants and their coaches in order to avoid any difference in exercise load among the participants. Football players who will train an average of 3 days a week and an average of 2 hours a day were included in the study. Both feet of the cases were evaluated separately. The structural properties of the foot have been studied. Patients with known systemic, inflammatory, neurological and degenerative diseases, those with lower back or lower extremity pain, those with lower extremity joint deformity, those with lower extremity operation history and those with using insoles were excluded from the study. Our work was presented to the ethics committee for approval and accepted with the ethics committee numbered 20478486-369 dated 19.11.2014.

### 2.3 Demographic Data

After recording the personal information of each individual such as age, position, medical history and family history, preferred and non-preferred foot and the weekly training programs were also questioned. Then the evaluation methods below were applied to each participant in the same way.

### 2.4 Clinical Assessment

#### 2.4.1 General Anthropometric Measurements

For general anthropometric measurements, the body weights (kilograms) and height (meters) of the cases (with a Charder MS-3400 height-weight meter) were measured and the body mass index (kilograms / square meter) was calculated. The foot numbers of the cases were measured with Heider Mass device.

#### 2.4.2 Sports Injury Detection Form

For all participants, a sports injury detection form was initially filled. Then, new injuries during the follow-up were recorded, too. Injuries were registered by the same physician. The subjects were able to freely consult this physician. The sports injury detection form to be used here was created using the injury detection form created by the Fédération Internationale de Football Association (FIFA) Injury Consensus Group.^4^

#### 2.4.3 Joint Hypermobility Assessment

There are controversies in the literature about whether joint hypermobility is a risk factor for injury development in athletes. We included hypermobility evaluation in our study in order to contribute to the literature on this matter. Beighton scoring was used to assess hypermobility. People with a total Beighton score of four points or more were considered hypermobile. Although Beighton’s diagnostic criteria are not including shoulder, hip, ankle, and foot, it is generally accepted because of its comprehensive, easy to apply, and symmetrical evaluation of joints.^11–13^

#### 2.4.4 Foot Posture Assessment

There are studies investigating the relationship between foot posture and injury development in athletes, but contradictory results have been reported between these studies. We aimed to determine the foot posture using FPI-6 scale and to investigate the relationship of these identified foot postures with the development of disability and to contribute to the literature. FPI-6 is a practical and reliable method of evaluating foot posture and its validity and reliability are proven.^14–16^ FPI-6 evaluation is performed when the patient is comfortable standing with both legs pressed. Both feet are evaluated separately. A total of 6 points on the front foot and the back foot are assigned a score of -2 to +2 according to their position, and as a result, all scores are evaluated to determine whether the foot is neutral, supine, or pronated. A value of -2 indicates an excess of supination for each point, while a value of +2 indicates for excessive pronation, and 0 indicates a neutral position.

The FPI-6 score was determined and recorded and each participant’s feet photographs simultaneously were taken. These scores were evaluated by two individuals and it was determined that there was no difference. First step of FPI-6 scale (assessment of the position of the forefoot relative to the back foot) was shown in Figure 1.

**Figure 1:**
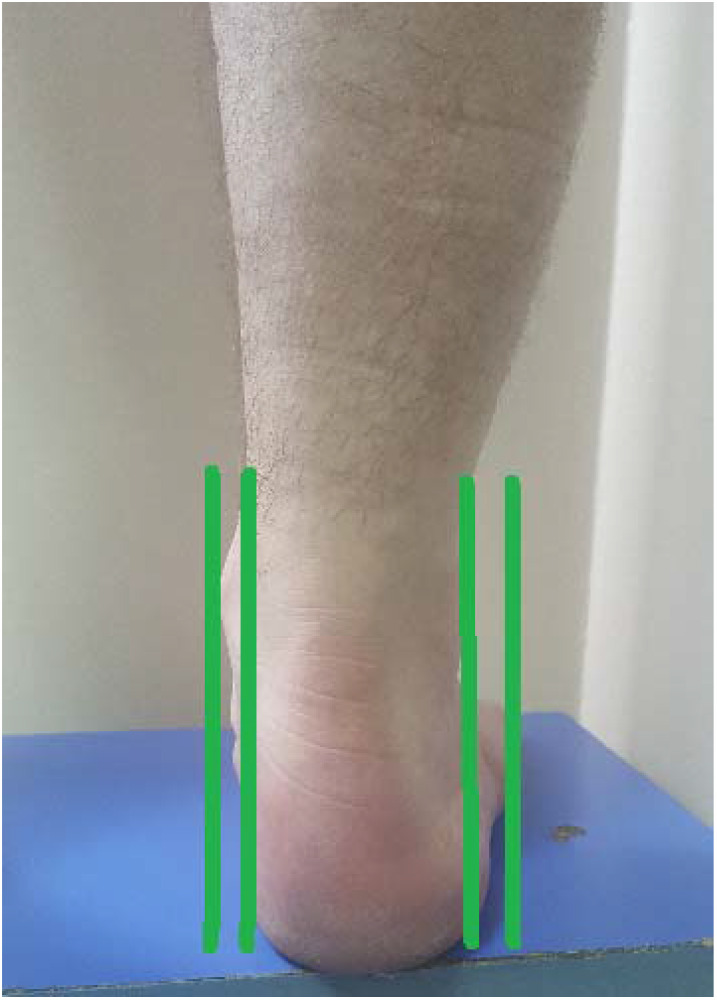
Assessment of the position of the forefoot relative to the back foot according to FPI-6

#### 2.4.5 Evaluation of Joint Range of Motion (ROM) with Inclinometer

There are studies in the literature investigating the relationship between ankle range of motion and development of disability.^17^ However, most of these studies focused on ankle dorsiflexion and plantarflexion. Few studies have investigated the relationship between ankle inversion and eversion measurements and the development of disability. We made four different joint range of motion measurements, including inversion and eversion ROM measurements of the ankle, which we think will contribute to the literature, and investigated their relationship to the development of disability. Measurements were made using (JTECH Medical Dualer IQ brand, USA) an inclinometer. Four movement capacities of each foot were recorded, separately for the right and left feet of the patients. Each measurement was made three times and averages recorded. Before the evaluation of the inclinometer, the subject was laid on his back on the examination table and was inspected using a right-angled wooden brace to reveal that both feet were 90 degrees in angle. This position was accepted as the starting point for the measurement of plantar and dorsal flexion movements. Before measurement, the inclinometer was reset to 90 degrees on the wall. For measurement, the probe of the inclinometer was placed on the footwell. First, dorsal and then plantar flexion movement widths were measured. Plantarflexion range of motion assesment was shown in Figure 2.

**Figure 2:**
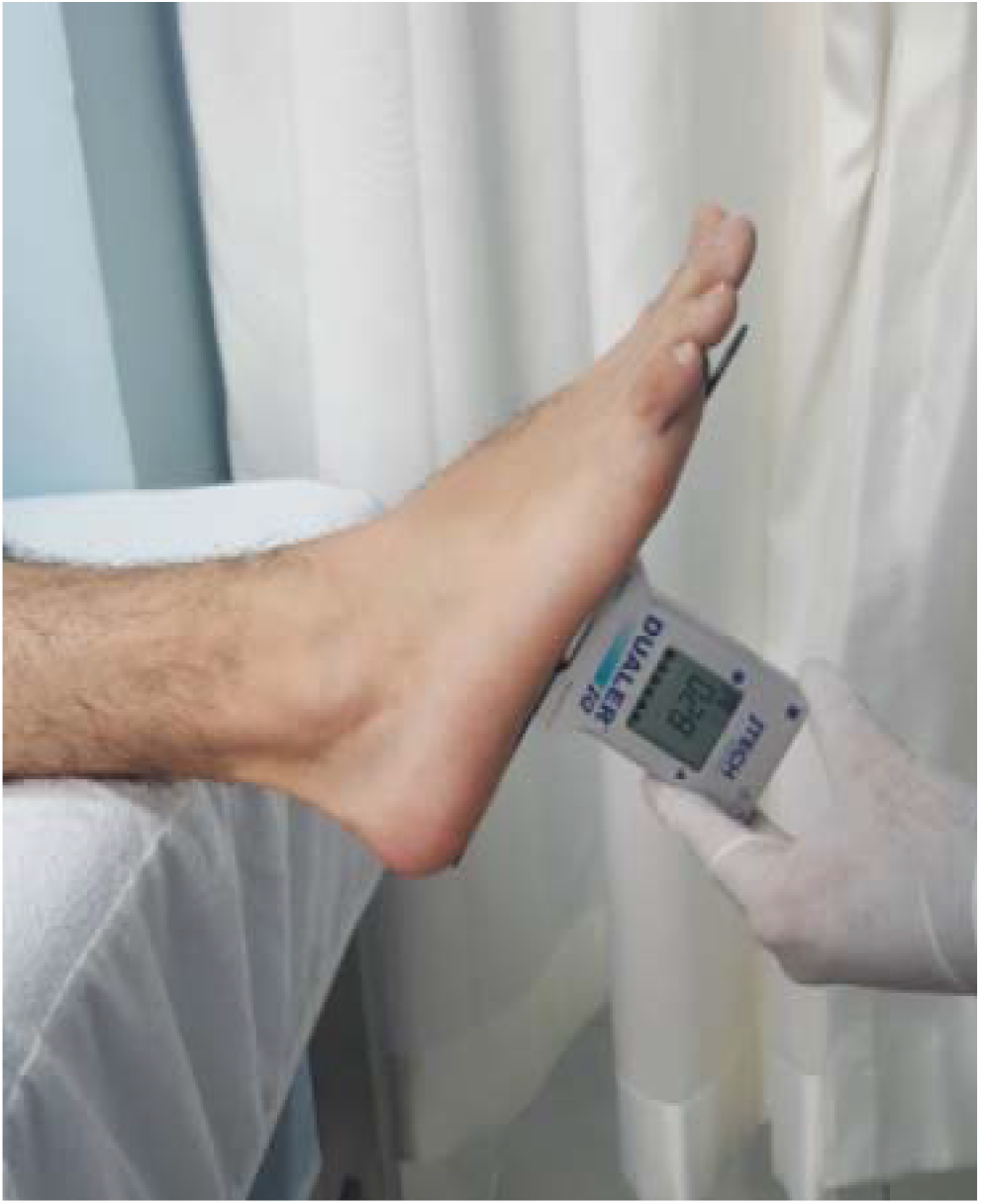
Plantarflexion range of motion assesment with inclinometer

For the measurement of inversion-eversion movements, the subject was placed on the examination table as aside. While both hips were in extension, the knee on the side of assessment was held in extension while the other knee was on flexion. During the measurement, the instrument was reset by placing the probe of the inclinometer so that it is perpendicular to the first metatarsal on the medial arc of the foot. The inversion movements were first measured, then the eversion movements were measured and the averages were noted.

#### 2.4.6 Evaluation of first Metatarsophalangeal Joint (MTP) Range of Motion

Although the first MTP joint is generally ignored during the examination, it has been reported in the literature that its evaluation will have important contributions to the physician in the examination of the foot.^18^ Participants’ first MTP joint dorsiflexion and plantarflexion values of both feet were measured 3 times with goniometer while participants were in the supine position. Then all measurements were averaged.

### 2.5 Plantar Pressure Evaluation (Pedobargraphic evaluation)

Foot loading patterns in football have been described in previous studies, and alterations in biomechanical loads have been suggested to be closely associated with foot and ankle overuse injuries.^8,10^ However, in static and dynamic pedobarographic measurements of the foot, data on the relationship between pressure values detected in various areas of the foot and disability development are contradictory. We aimed to determine the relationship between the values we detect and the development of disability by making static and dynamic pedobarographic measurements and to contribute to the literature in this way.

Measurements of the foot plantar pressures of the cases were made with a pedobarograph device (RsScan International pedobarograph device, 1m, 3D Scientific + Balance software model, Belgium). This system measures the pressure of the foot base as static (standing) and dynamic (walking). The pressure measuring platform of the device has a 975×325 mm sensor area within a general frame of size 1068×418×12 mm, with a total of 8192 sensors, with 4 sensors per cm^2^ falling. The frequency is 500 Hz, the pressure range is 0-200 N / cm^2^, the temperature range is 15°-40° C, connection power is 220/110 volts.

When making a static assessment, the attention of the cases was disseminated with the questions asked so that they could not direct bodyweight to a particular side of the platform. For a balanced evaluation, they were asked to look at a fixed point on the wall. While standing on the platform, the average distance between the two feet was determined as 8 cm. The evaluation was made at the same time for both feet. Maximal pressure measurements were evaluated in seven different areas (heel-medial, heel-lateral, midfoot, forefoot-medial, forefoot-middle, forefoot-lateral, fingers) in N / cm^2^. Pedobarographic static foot plantar pressure evaluation was shown in Figure 3.

**Figure 3:**
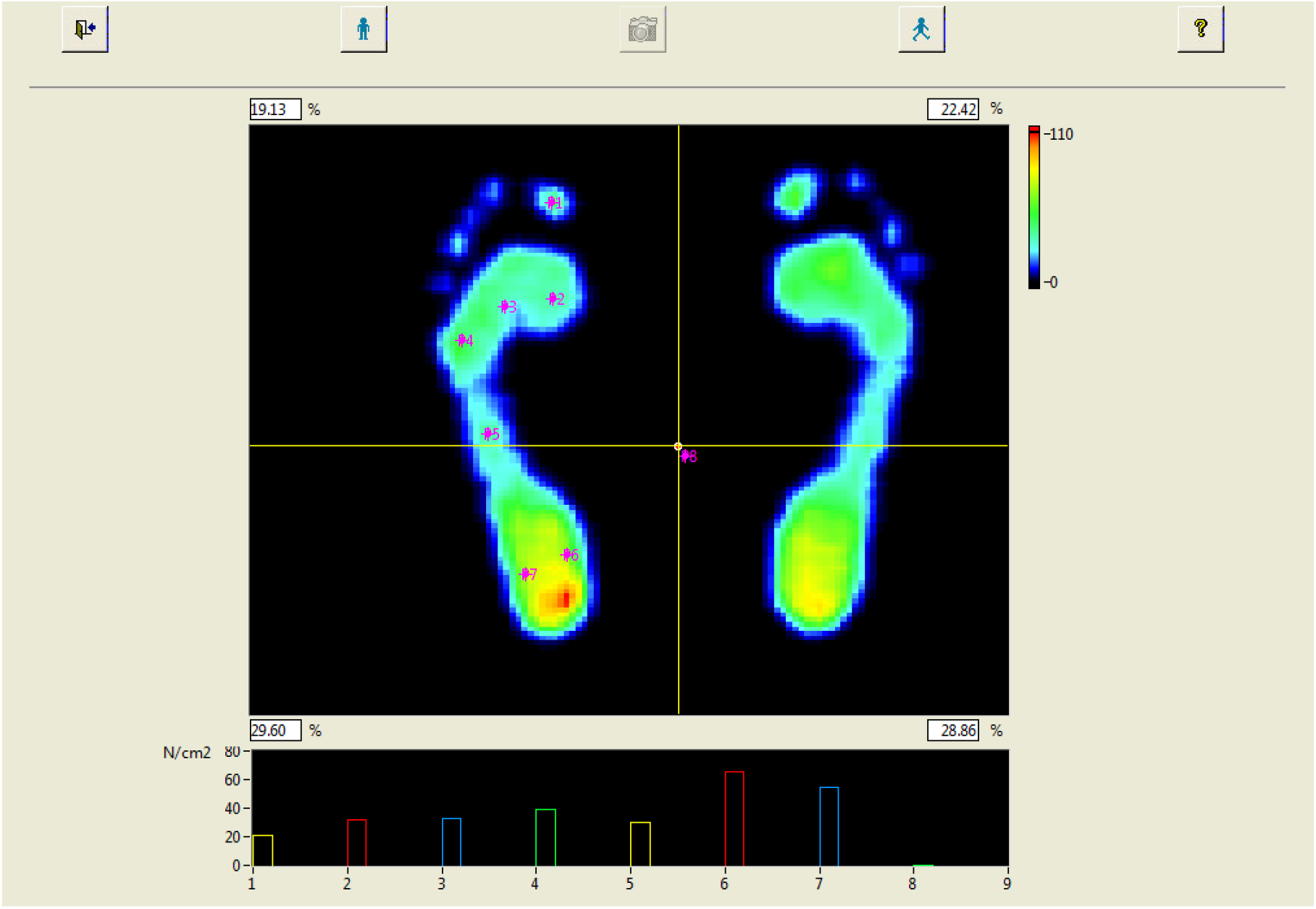
Pedobarographic static foot plantar pressure evaluation

For dynamic evaluation, subjects firstly asked to walk on a 30-meter-long field for a few minutes to generate normal walking speed. Then they were asked to continue their normal walk and press the pressure platform with a normal step. The subjects were asked to walk again in the situation of standing still on the platform or pressing the platform incorrectly. Measurements were made 10 times for both feet. As dynamic pedobarographic analyzes, maximum dynamic pressure assessments, and load velocity assessments were performed in ten different areas (heel-medial, heel-lateral, middle foot, forefoot, five metatarsal regions, thumb, and other fingers).

### 2.6 Follow-up Evaluation

After the initial evaluation, the participants were followed for 12 months in terms of new foot and ankle overuse injury development. Overuse injury was defined as “one caused by repeated micro-trauma without a single, identifiable event responsible for the injury”.^19^ The type of injury and the injured body region were recorded.

It was planned to diagnose and record the injuries by the same physician. The subjects were able to freely consult this physician. Evaluation and registration of the injured football players were made by the same physician. During the follow-up, four footballers were excluded because they did not meet the inclusion criteria. The clinical and pedobarographical evaluations of football players with at least one foot and ankle overuse injury at twelfth month and those who did not have any foot and ankle overuse injury were compared between the two groups.

### 2.7 Statistical Analyzes

Power analysis was performed before the study and the size of the sample according to the power analysis result is calculated as the minimum of 100 persons for α*= .05 d= .40 (d = effect size)* and for 80% power.

SPSS 21.0 package program was used for the statistical analysis of the data. Mean and standard deviation (SD) values were calculated as descriptors for the numerical data. The Mann-Whitney U test was used to compare independent groups, and the Wilcoxon sign test was used to compare dependent groups when the number of individuals in the groups was less than 30. Independent samples t-test was used to compare independent groups and a paired t-test was used to compare dependent groups when the number of people in groups was 30 or more. Statistically, significant significance was accepted as *p<.05*. Pearson correlation analysis was used to investigate the correlation of the data.

## 3. FINDINGS

The average age of 100 participants was 20 (range, 16-33), the average height was 177 centimeter (range, 164-192 cm), the average weight was 71 kilogram (range, 57-86 kg) and the average BMI was 23 (range, 18-27).

When the distribution rates of 100 footballers according to their positions were examined, it was seen that the central midfielders (n: 27) followed by the centre-backs (n: 19) were the most. Distribution rates of all positions are given in Table 1.

**Table 1:** Distribution rates of 100 footballers according to their positions

| Position | n | % |
| --- | --- | --- |
| Goalkeeper | 9 | 9,00 |
| Centre-Backs | 19 | 19,00 |
| Central Midfielders | 27 | 27,00 |
| Fullbacks | 17 | 17,00 |
| Wide Midfielders | 14 | 14,00 |
| Attacking Midfielders / Strikers | 14 | 14,00 |
| Total | 100 | 100,00 |

Footballers’ Beighton scores, Foot Posture Index-6 scores, ankle joint ROM values measured by digital inclinometer, and first MTF joint ROM values measured by goniometer were analyzed. These values are shown in Table 2.

**Table 2:** Participants’ Beighton Scores, Foot Posture Index (FPI-6) Scores, Joint Range of Motion Values

|  | n | Minimum | Maximum | Mean | SD |
| --- | --- | --- | --- | --- | --- |
| Beighton Score | 100 | 0,00 | 6,00 | 1,42 | 1,48 |
| FPI-6 Right Foot | 100 | -1,00 | 4,00 | 0,88 | 1,14 |
| FPI-6 Left Foot | 100 | -3,00 | 3,00 | 0,68 | 1,13 |
| Ankle Dorsiflexion Right Foot | 100 | 8,00 | 25,00 | 17,82 | 3,24 |
| Ankle Dorsiflexion Left Foot | 100 | 10,00 | 25,00 | 18,00 | 3,40 |
| Ankle Plantarflexion Right Foot | 100 | 30,00 | 62,00 | 45,33 | 6,23 |
| Ankle Plantarflexion Left Foot | 100 | 30,00 | 63,00 | 45,27 | 6,31 |
| Ankle Inversion Right Foot | 100 | 20,00 | 52,00 | 35,46 | 5,43 |
| Ankle Inversion Left Foot | 100 | 18,00 | 60,00 | 34,04 | 5,71 |
| Ankle Eversion Right Foot | 100 | 8,00 | 28,00 | 16,98 | 4,38 |
| Ankle Eversion Left Foot | 100 | 5,00 | 31,00 | 16,19 | 4,44 |
| First MTP Dorsiflexion Right Foot | 100 | 20,00 | 66,00 | 44,56 | 7,61 |
| First MTP Dorsiflexion Left Foot | 100 | 20,00 | 60,00 | 45,59 | 6,67 |
| First MTP Flexion Right Foot | 100 | 15,00 | 50,00 | 38,86 | 7,97 |
| First MTP Flexion Left Foot | 100 | 15,00 | 50,00 | 38,13 | 8,00 |

Generalized joint hypermobility conditions of the players were evaluated according to the Beighton Score. Participants with a Beighton Score of 4 or above were considered to have generalized joint hypermobility. While 88 of the participants had no generalized joint hypermobility, this was present in 12.

In the follow-up of the players, at the end of the twelfth month, when 31 of the 100 football players who participated in the study had an overuse-type injury, and 3 participants had 2 injuries. The total number of injuries occurred was 37.

Metatarsalgia (n:17) was the most common type of injury. It was followed by plantar fasciitis, achilles tendinopathy, and metatarsal stress fracture, respectively. Injury types of football players who experienced injury within 12 months are shown in table 3.

**Table 3:**
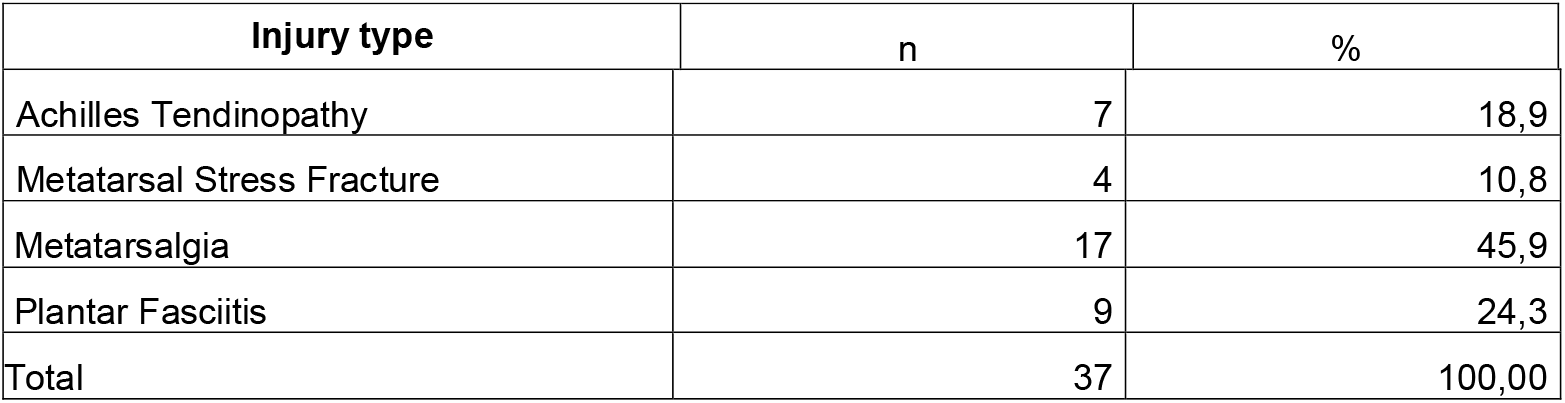
Overuse Injury Types of Football Players

The pedobarographic plantar static pressure values of the group who had at least one injury and the group without any injury were compared using the T-test at the end of the twelfth month. Static forefoot mid-zone pressures of non-preferred foot were significantly higher in the group with at least one injury compared to the group without injury *(p = .040).* In other pressure regions, no statistical difference was found between the two groups.

The pedobarographic maximum dynamic pressure values of the group who had at least one injury and the group that did not have any injury were compared using the T-test at the end of the twelfth month. There was no statistical difference in the dynamic pressure values between the two groups.

The pedobarographic load rate values of the group who had at least one injury at the end of the twelfth month and the group without any injury were compared using the T-test. There was no statistical difference in load speed values between the two groups.

The ankle and first MTP joint range of motion values of the group that had at least one injury and the group that did not have any injury were compared using the T-test. In the group with at least one injury, non-preferred side ankle plantarflexion, preferred side ankle eversion, and non-preferred side first MTP joint dorsiflexion ROM values were significantly decreased compared to the group without injury *(p = .044, p = .029, p = .023, respectively).* There was no statistically significant difference in other findings.

Foot postures determined according to the FPI-6 scale of the footballers at the end of the 12-month follow-up were compared between the group with at least one injury and the group without any injury. In the evaluation for the preferred foot, 3 of 11 footballers with supine foot posture developed an injury, while 31 (34.8%) of 89 football players with neutral foot posture developed an injury. To determine whether there is a statistical increase in the risk of injury in players with a supine foot posture, the injury rates between footballers with a supine foot posture and those with a neutral foot posture were compared for the preferred and non-preferred foot using the Pearson chi-square test. There was no statistically significant difference between the two groups *(p: .448, p: .226, respectively).*

## 4. DISCUSSION

This study consisted of 100 licensed football players between the ages of 16-33 who played football in the A2 team, which is the second team of the clubs, who regularly train and participate in football competitions in Izmir and Manisa provinces. It was aimed to determine the structure of foot and ankle of the players with clinical examination and FPI-6 scale, to reveal the ROM values of the ankle and first MTP joint, to determine the pedobarographic static and dynamic pressures and loading rates of the foot and to determine the hypermobility status of the players according to the Beighton Score. Then, the players who had an overuse injury at foot and ankle region were identified in the follow-up, so the clinical and laboratory values of the injury and non-injury group were compared and the risk factors that might prepare the ground for the occurrence of overuse injury were determined.

Football is the most popular sport in the World. FIFA (Fédération Internationale de Football Association) has approximately 250 million licensed players from 204 countries. About 1% of these players are professionals.^20,21^ Football requires high coordination and skill of the body, especially the lower limbs. It is known that body structure is important for sports performance. The foot structure is important in functions such as standing, walking, running, and jumping in many sports branches and is effective in the performance and success of the athlete.^22,23^

The foot is easy to examine but difficult to diagnose because of its complex functional anatomy. Disturbances in the structure and function of the foot can lead to prominent restrictions on the activity and performance level of the athlete. It is thought that the risk factors that may cause injury can be shown by revealing the soles of the foot and the corrections to be made can contribute to the improvement of sports performance.^24,25^ The full functioning of the foot is not only important in terms of foot health, but it is also necessary for the health of our entire body, especially neighboring joints.^1,3^ Foot pressures caused by unbalanced anatomical positions are the cause of physiological disorders in the body and musculoskeletal system. These repetitive abnormal effects can directly affect the ankle and structures adjacent to the ankle, even vertebral colon diseases. A direct relationship was found between pressure distribution disorders in various parts of the sole and an increase in scoliosis level.^26^

Foot loading patterns in football have been described in previous studies, and alterations in biomechanical loads have been suggested to be closely associated with foot and ankle overuse injuries. A recent study assessing plantar pressure under static conditions observed larger pressure values for the nonpreferred foot in young soccer players than in the control group.^1^ Various foot and ankle problems result from repetitive high loads on the foot.^8^ It has also been reported that stress fracture is common among soccer players, and that 38% (9/24) of players on the US team developed stress fractures during the 1994 FIFA World Cup.^10^

Reports on the association between static foot plantar pressure and injuries are contradictory. In their prospective cohort study, Brund et al.^27^ reported that runners displaying a more medial pressure during stance phase at baseline sustained a greater amount of foot and ankle overuse injuries compared to those displaying a lateral shod pressure during stance phase.

In a study by Azevedo et al.^1^, they found that asymmetry between foot soles pressures in football players is associated with stress injuries. The foot pressure of 15 young footballers and the control group of 15 people was measured with the Matscan (Tekscan, Inc., Boston, USA) device while the footballers were standing barefoot on the platform and higher pressure values were determined in the non-dominant standing thumb, fifth metatarsal region and heel medial.

We found asymmetric pressure distribution between the preferred and non-preferred foot in the group who had an injury in the pedobarographic static foot plantar pressure measurements. We compared the foot static pedobarographic maximum pressure values of the football players who had an injury with the static maximum pressure values of the group who did not have any inury during 12 months of follow-up. We detected increased pressures in the anteromedial area of non-preferred foot *(p = .040).* We think that, asymmetry in foot plantar pressure distribution between preferred and non-preferred foot and pressure elevations detected in forefoot in static measurements may be a predisposing factor for injury formation in football players. Relationships between injury history and static plantar pressure measurements are shown in Table 4.

**Table 4:**
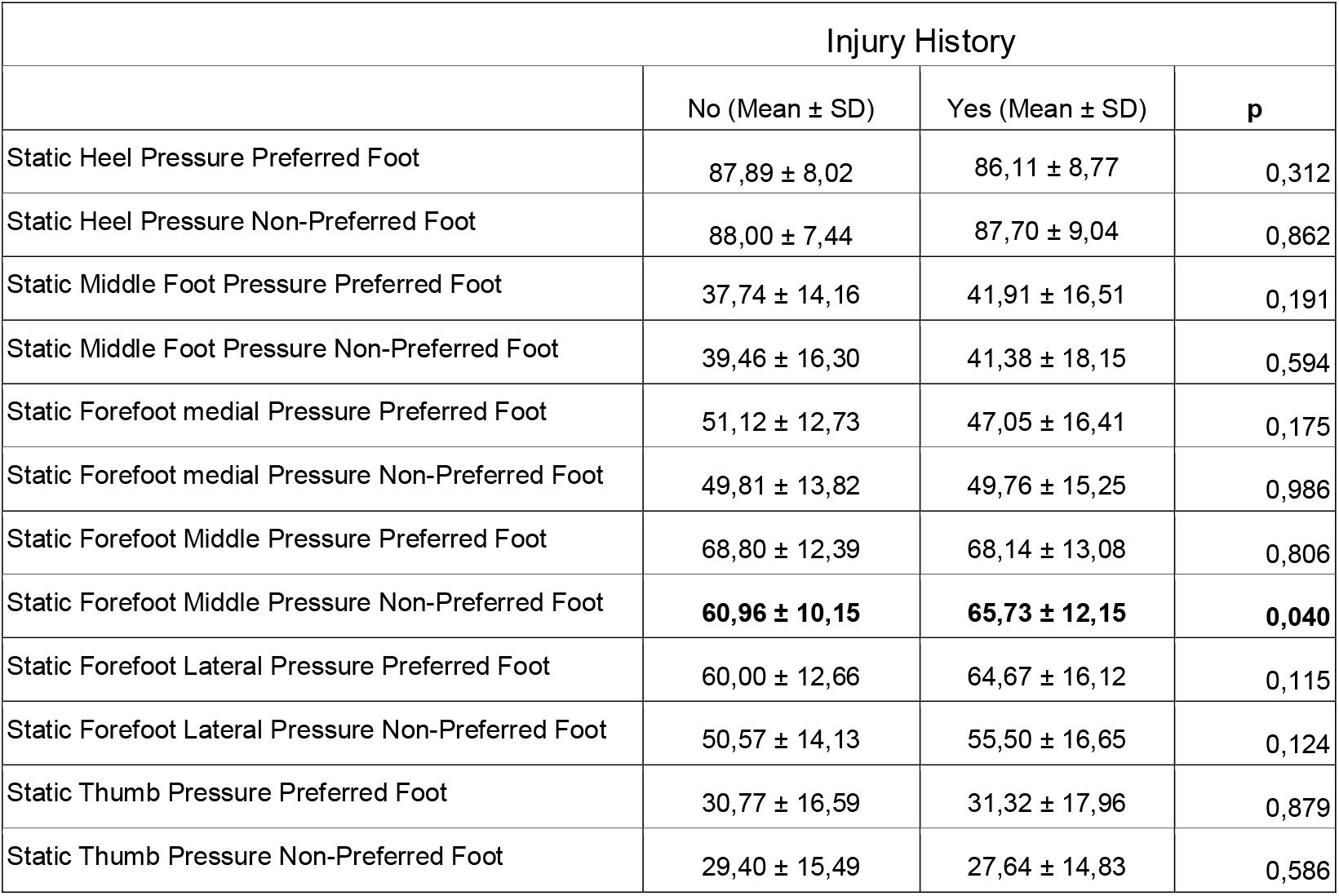
Relationship between Injury History and Static Plantar Pressure Measurements

Wong et al.^9^ investigated the relationship between the movements performed during the football game and the soles of the feet and determined high-pressure values on the medial side of the foot. They interpreted this situation as a condition that would predispose to injury. 15 male football players (mean age 20.90 ± 1.3) were included in the study and recorded with 99 sensor pressure recording devices placed in the soccer shoe and these pressures were recorded by dividing the foot into 10 regions. The highest pressure values were recorded in the thumb and heel medial and it was thought that these high-pressure values may cause a decrease in the comfort of the foot and chronic injury.

Willems et al.^28^ examined possible risk factors for exercise-related lower leg pain (ERLLP) related to walking and running style in 400 physical education students (Age range: 17-28, mean age: 18.4). They determined the jogging and walking structures of 400 students at the beginning of the academic semester through plantar pressure measurement and 3D walking kinematics. After these measurements, all sports injuries occurring throughout the study were recorded by the same sports physician and followed for 26 weeks by giving a similar sports program. Compared to the pre-injury controls of patients with ERLLP, they found an increase in pressure at the medial edge of the foot, a decrease in pressure at the lateral edge, and increased pronation.

We compared the data obtained from the pedobarographic measurement of foot plantar maximum dynamic pressures of football players who had at least one injury in 12 months of follow-up with the foot plantar maximum dynamic pressure data of football players who never had an injury. There was no statistical relationship between the injuries and the dynamic plantar pressure measurements. Unlike the studies we mentioned in the literature, we could not find a statistically significant relationship between dynamic plantar pressure measurements and injury development.

Whether hypermobility is a risk factor for disability in athletes is controversial and there are studies in the literature that provide an opposing opinion on this issue. Smith et al.^29^ investigated the incidence of hypermobility in young female netball players and the relationship between hypermobility and injuries; and hypermobility was significantly associated with increased injury prevalence in young netball players. On the other hand, Richard et al.^30^ studied the relationship between generalized joint hypermobility and injury in the study of 33 male professional football players playing in England second League, and found the injury rate similar between groups with and without hypermobility.

In our study, all football players were evaluated in terms of joint hypermobility syndrome (JHS) with Beighton scoring. According to the Beighton score, the players scoring four and above are considered to be hypermobile. JHS was detected in 12 players, while JHS was not detected in 88 players. It was observed that 25% of the footballers who had JHS had an injury, and 25% of the players without JHS had an injury in the evaluation we made in the twelfth month. Whether hypermobility is a risk factor for injury, when compared between the two groups using Pearson chi-square test, there was no significant difference in terms of hypermobility between the two groups. *(p = .140, p = .653).*

FPI-6 is a practical and reliable evaluation method that is used to evaluate foot posture, validity, and reliability.^16^ With the FPI-6 used in our study, the total score is obtained between -12 and +12 as a result of the evaluation made by looking at the 6 different points of the foot. It shows the supination posture from 0 to -12 and the pronation posture from 0 to +12. In the literature, studies have been carried out to determine the normal values of FPI-6. In the study conducted by Redmond et al.^31^, the FPI-6 scores were categorized and reported that the scores between -12 and -5 were highly supine feet, values between -4 and -1 were supine feet, values between 0 and +5 were neutral feet and they reported that values between +6 and +9 reflect prone to foot, and values between +10 and +12 reflect a high degree of prone foot posture. In the literature, different score ranges have been reported in various studies (14,29,31,32) regarding the cut-off values of the FPI-6 scale to be used in supine, neutral, and prone foot categorization. We used the cut-off values determined by Redmond et al. in our study.

Cain et al.^32^ found that there was a statistically significant risk factor for foot/ankle overuse injuries in adolescent football players with mild prone (FPI-6 score +2 and below) and supine foot posture in their studies of 76 adolescent male football players using the FPI-6 assessment scale.

100 players were included in our study and FPI-6 scores were examined and recorded separately for both feet at the start of the study. Afterward, we compared the FPI-6 scores of the players who had at least one injury in the twelfth month with the group who never had any injury. We did not find a statistically significant relationship between the group who suffered from an injury and the group that did not have any injury, to determine the predictive value of the FPI-6 score for injury, using the Pearson chi-square test for the right and left foot separately *(p> .05).*

However, in our study, we did not detect any moderate or severe prone foot type with FPI-6. As one of the possible reasons for this, we thought that people with moderate to high prone foot structure would not be in this population since they would have difficulty doing a sport that requires speed, balance, and condition like football. We think that different cut-off values should be determined for relatively healthy, active sports people with a low foot arch to define the foot posture classification. We think that more extensive further studies are needed for this population.

Cerrahoglu et al.^17^ reported that the restriction of joint movements in the foot causes mechanical stress and therefore, it is predisposed to increase in plantar pressure and foot/ankle injuries. Although the first MTP joint is generally ignored during the examination, it has been reported in the literature that its evaluation will have important contributions to the physician in the examination of the foot.^18^

Plantar fasciitis is the most common diagnosis (10% to 15%) for patients with foot and ankle pain. Plantar fasciitis has a multifactorial etiology. It was previously thought to be an inflammatory syndrome; however, recent studies have emphasized that a degenerative process is more dominant. In their study, Ulusoy et al. described the factors thought to be associated with the disease include biomechanical dysfunction, mechanical overload, obesity, overuse, achilles tendon strain, decreased ankle dorsiflexion, atrophy of the intrinsic muscles, and a pronated foot type.^33^

In the study conducted by Halabchi et al.^22^ for intrinsic risk factors that will predispose ankle injuries, 58 footballers were included in the study, and the relationship between ankle ROM and injury was investigated. They reported that the limitation of left foot plantarflexion was statistically significantly associated with acute and chronic ankle injuries in athletes. They did not find the same significant relationship in the ankle dorsiflexion.

When we compared the ROM values of footballers who had an injury and footballers who did not have an injury, we found a significant limitation in the group that suffered an injury, in the ankle eversion, first MTP joint dorsiflexion and ankle plantarflexion degrees *(p = .029, p = .023, p=.044, respectively)*. Injury and joint ROM relationships are shown in Table 5.

**Table 5:** Injury - Joint Range of Motion Relationship

| Injury History |  |  |  |
| --- | --- | --- | --- |
|  | No (Mean ± SD) | Yes (Mean ± SD) | p |
| Preferred Foot Ankle Dorsiflexion | 17,78 ± 3,38 | 18,08 ± 2,02 | 0,482 |
| Non-Preferred Foot Ankle Dorsiflexion | 18,11 ± 3,39 | 17,16 ± 3,48 | 0,302 |
| Preferred Foot Ankle Plantarflexion | 45,15 ± 6,21 | 46,58 ± 6,47 | 0,195 |
| Non-Preferred Foot Ankle Plantarflexion | **46,18 ± 6,02** | **43,50 ± 6,57** | **0,044** |
| Preferred Foot Ankle Inversion | 35,56 ± 5,61 | 34,66 ± 4,00 | 0,298 |
| Non-Preferred Foot Ankle Inversion | 34,26 ± 5,90 | 32,41 ± 3,84 | 0,117 |
| Preferred Foot Ankle Eversion | **17,26 ± 4,49** | **14,91 ± 2,84** | **0,029** |
| Non-Preferred Foot Ankle Eversion | 16,30 ± 4,56 | 15,33 ± 3,47 | 0,242 |
| Preferred Foot MTP Dorsiflexion | 44,31 ± 7,98 | 46,33 ± 3,74 | 0,219 |
| Non-Preferred Foot MTP Dorsiflexion | **48,33 ± 3,89** | **45,21 ± 6,89** | **0,023** |
| Preferred Foot MTP Flexion | 39,07 ± 8,26 | 37,25 ± 5,37 | 0,207 |
| Non-Preferred Foot MTP Flexion | 38,27 ± 8,13 | 37,08 ± 7,21 | 0,190 |

In parallel with the literature, we think that ankle plantarflexion and first MTP joint ROM limitations are associated with foot/ankle overuse injuries. There are few studies in the literature about the relationship between ankle inversion and eversion limitations and injury. In our study, we found a significant relationship between ankle eversion restriction and the risk of developing an injury. We think that ankle and first MTP joint ROM limitations cause mechanical stress in the foot, disrupt foot biomechanics, and thus cause susceptibility to foot/ankle injuries. We believe that ankle and first MTP joint ROM examination in football players is important in terms of revealing an underlying ROM limitation and preventing possible future injuries by taking precautions for this.

The loading speed or time pressure integral (N / cm^2^ sec) is a parameter that can be measured pedobarographically, which is defined as the amount of time that must pass before the evaluated foot region reaches maximum pressure. It is an expected and necessary situation in footballers that the loading speed is lower than the general population. This can be explained by the rapid displacement of the footballers and the reaction times of the foot soles from the contacted surfaces quickly due to the minimal reaction times.

When we examined the literature, we found that studies about the pedobarographic loading rate in the literature are few. Uzun et al.^34^ evaluated the loading speed in their study with 22 female footballers (18,80 ± 2,2 years) and 28 female volunteers (27,5 ± 6,6 years) as a control group. They found that there were significant differences in loading speed values between the female footballer and the control group in the second and third metatarsal heads of the right foot and the heel medial, heel lateral, and in the second, third and fourth metatarsal heads of the left foot. In our study, we evaluated the pedobarographic foot loading speeds of footballers from 10 different parts of the foot (thumb, second-fifth fingers, five metatarsal regions, middle foot, heel medial, and lateral). When we compared the foot loading speed values of the footballers who had an injury in the follow-up and the footballers who had no injury, to determine the relationship between the foot loading speed values of the participants with the injury, we did not find a statistical difference between the two groups *(p> .05).*

### Study Limitations

A limitation of this study is the fact that the subjects underwent plantar pressure measurements during barefoot walking and the foot–shoe–ground interface was not taken into account (as running shoes might correct or even exacerbate plantar pressure distributions). Nevertheless, we choosed barefoot instead of with-shoe walking because this condition permits focusing on intrinsic foot biomechanics without interference of different or unusual footwear. Willems et al.^28^ showed that intrinsic plantar pressure risk factors for ERLLP were less pronounced in shod compared with barefoot running. Therefore, those authors advised the use of barefoot running to be implemented first.

## 5. CONCLUSION

The main purpose of this study was to compare the values determined in the clinical examination of the foot and ankle and the plantar pressure measurements of the foot in athletes who developed an overuse-type disability and in athletes who did not develop overuse-type disability, and was to find factors that might predispose to disability, during the one-year follow-up. If we summarize what we found from these parameters;

I. We found that asymmetry in foot plantar pressure distribution and pressure elevations detected in anteromedial regions of foot in static measurements, may be a predisposing factor for foot and ankle overuse injury formation in football players.
II. In parallel with the literature, we found that ankle plantarflexion and first MTP joint ROM limitations are associated with foot and ankle overuse injuries. Also, we found a significant relationship between ankle eversion restriction and the risk of developing an injury. We believe that ankle and first MTP joint ROM examination in football players is important in terms of revealing an underlying ROM limitation and preventing possible future injuries by taking precautions for this.
III. When we compared the injury rates of the group with and without hypermobility, we did not find a significant difference between the two groups. We do not think that hypermobility is a risk factor for injury development.

## 6. PERSPECTIVE

These findings suggest that limitations in ankle and foot ROM and impairments in foot plantar pressure distribution may be risk factors for the development of an overuse injury. For this reason, we recommend that early and routine joint ROM evaluation and plantar pressure evaluation be included in footballers’ examinations. We believe that correcting foot and ankle abnormalities that may occur with these evaluations can be effective in preventing possible overuse injuries and further studies are needed on this subject.

## Data Availability

all data is available

## ACKNOWLEDGEMENTS

All authors declare that they have no conflicts of interest. The authors also declare that this study has received no financial support from any person or organization.

